# Modeling Supraventricular Tachycardia Using Dynamic Computer-Generated Left Atrium

**DOI:** 10.1101/2023.03.28.23287860

**Authors:** Bryant Wyatt, Avery Campbell, Gavin McIntosh, Melanie Little, Brandon Wyatt

## Abstract

Supraventricular Tachycardia (SVT) is a type of irregular heartbeat seen when the heart’s upper chambers beat either too quickly or out of rhythm with the heart’s lower chambers. The loss of synchronization between the upper and lower chambers will result in perturbations of, blood flow. This is why SVT, which includes atrial fibrillation and atrial flutter, is a leading cause of strokes, heart attacks, and heart failure in the world today. The most successful treatment for SVT is catheter ablation, a procedure in which an electrophysiologist (EP) maps the heart to find areas with abnormal electrical activity. The EP then runs a catheter into the heart to ablate the abnormal areas, blocking the electrical signals or destroying the myocytes causing them. Not much is known about what triggers SVT and much research is still being done to find effective ablation strategies for various forms of SVT. We have produced a dynamic model of the left atrium accelerated on NVIDIA GPUs. An interface allows researchers to insert ectopic signals into the simulated atrium and ablate sections of the atrium allowing them to rapidly gain insight into what causes SVT and how to terminate them.

## INTRODUCTION

Heart disease and strokes are the leading causes of death worldwide.^1–4^ Supraventricular Tachycardia (SVT) is one of the most significant contributing factors contributing to strokes, heart failure, and in some cases, acute myocardial infarction.^18–20^ Hence, it is imperative that if we are to build healthier lives free of cardiovascular diseases and strokes, we need to reduce the occurrence of SVT. SVT encompasses all cardiac arrhythmias in which the underlying mechanism sustaining the abnormal heartbeat originates above the ventricles.^3,5^ This abnormal heartbeat can alter the natural synchronization between the atria and the ventricles, perturbing the laminar flow of blood through the heart, and causing it to stagnate in the small finger-like appendage of the left atrium. This, in turn, allows for the formation of certain types of lethal blood clots known as mural thrombi which can become dislodged and travel to the brain or coronary arteries, resulting in stroke or heart attack. It is this mechanism that causes individuals with atrial fibrillation (AF) to experience a five-fold increased risk of stroke.^3,6,7^

In a normally functioning heart, the sinus node is the pacemaker; it consistently produces an electrical impulse to rhythmically dictate the heart’s rhythm as well as rate. This electrical impulse causes a chain reaction that propagates throughout the heart, producing a life-sustaining heartbeat. Ectopic electrical impulses can cause chain reactions to occur at the wrong place and at the wrong time, disrupting the normal sinus rhythm. This can cause the atria to flutter, beat out of sync with the ventricles, or have a myriad of other undesirable presentations. The beating heart is a multi-dimensional nonlinear dynamical system that is sensitive to initial conditions. Hence, SVT can produce chaotic outcomes that are impossible to predict with great accuracy ^2,5,8,9^

There are two basic mechanisms of action that can cause SVT.^2,10,11^ The simplest of the two is focal tachycardia. Focal tachycardias are caused by increased automaticity in specific regions of the heart overriding the sinus node, causing the entire heart to mistakenly follow its lead.^12–14^ The second form of SVT is called reentry tachycardia and is a far more complex system. Reentrant tachycardias involve intricate multivariate feedback loops that can be unique to individual anatomy and local electrical properties.^3,10,14–17^ While many forms of reentry can be explained by anomalous anatomical features such as extra electrical connections between what should be electrically isolated areas of the heart, other circuits remain poorly understood or can result from previous ablation lesions.^10,11,16,17^ AF, which many experts feel should be in a category of its own, is the least understood of the tachycardias and most prevalent in patients.^1,5,15,18^ AF is a chaotic arrhythmia that takes over the heart. It is difficult to determine what triggers AF and equally as hard to know where to place ablation lesions to eliminate AF.^1,15^ Chaos is studied in an area of mathematics known as Dynamical Systems and was made popular by Edward Lorenz in his study of weather prediction.^19^ AF and the weather are similar in that they are both sensitive to initial conditions. Meaning that small changes in their initial state can cause dramatic results in final outcomes, popularly known as the butterfly effect. Such systems are impossible to solve analytically and must be studied numerically, using computer models.^20^ Many physicians in the electrophysiology community hold competing theories explaining a particular subset of SVT.^1,15,21^ Although the explanatory validity of many of these theories is mutually exclusive, each will still develop a seemingly effective ablation strategy.^1,21–25^ This curiosity stems from the fact that the heart is a nonlinear dynamical system, and why this problem lends itself well to being attacked with computational mathematics.^20^

In many cases, SVT can be successfully controlled with medication and lifestyle changes. However, many of these drugs are difficult for patients to tolerate, and some medications with the most established efficacy are known to be hepatotoxic and have many deleterious side effects.^24,24,26,27^ Hence, catheter ablation, though more invasive than medication therapy, has proven to be the most reliably efficacious and safest method physicians have in treating patients with recurring SVT.^4,22–24,24,28,29^

Catheter ablation is a minimally invasive procedure that requires an electrophysiologist (EP) to build a three-dimensional electro-anatomical map of the heart to aid in identifying areas of abnormal electrical activity.^2,4,22,30^ The EP then runs an ablation catheter into the heart and using the computer map displayed on the screen, ablates either the areas from where abnormal electrical activity originates or electrically isolates those areas so they cannot propagate through to the rest of the heart.^4,22,30^

Radiofrequency (RF) catheter ablation and three-dimensional electro-anatomical mapping techniques have seen dramatic improvement over the last 10 years, allowing doctors to perform procedures on beating hearts that until recently were thought impossible.^22,24^ But, much is still not understood about what causes heart arrhythmias and how to use RF catheter ablation to treat them.^23,24,26,28,31^ In many cases, patients that have undergone ablation procedures will have to return for additional treatments.^28,32^ The heart is a nonlinear dynamical system, which makes it difficult to predict the precise outcome that will result from changes to the system, such as RF catheter ablation lesions.^33^ What would help doctors and researchers is a way to test out ideas rapidly and inexpensively.^8,22,24,29^ This could be accomplished with an interactive dynamical model of the heart. Such a model will improve clinical decisions leading to better patient outcomes and helping make a world where people can live longer healthier lives.

Electrical modeling of a heart has been done.^34^ Researchers lack a computer-generated model of a beating heart on which to perform experiments. We have created an n-body model of the left atrium that beats in real-time and is adjustable down to the individual muscle level.^33^ The interactive model allows doctors and researchers to study arrhythmias and RF ablation treatment plans outside of the operating room. The left atrium was chosen because it is where most complex arrhythmias arise.^1,2,18^

## METHODS

### Cardiac Muscle Simulation

The cardiac muscles are modeled as adjustable Hookean springs that smoothly transition between two states: the resting state and the contracting state.^33,35^ The cardiac tissue is modeled as a network of muscles and connecting nodes. Each muscle is stored in a dynamic structure that keeps track of the muscle’s resting strength and length, contracting strength and length, electrical wave propagation speed, and initiation end of electrical stimulation.^33,36^ Each muscle also contains an internal clock that keeps track of how long the muscle is in its contraction phase, how long it is in its recharge phase, and the position of the electrical wavefront as it propagates down the muscle.^36^ Each connecting node is also stored in a dynamic structure. This structure keeps track of every muscle that is connected to the node, and if the node is ablated (turned off).^11,35^

If an unablated node is hit by an electrical wavefront, it will try and excite every muscle in its connection list.^9,11^ If the node is ablated, it will simply ignore the signal. If a muscle is excited and in its rested state, it will record which end of the muscle received the impulse, reset its internal clock, put the muscle into its contracting state, and start an electrical wavefront propagating in the proper direction. If the muscle is in its recharge state or if the muscle is already in a contracting state, the muscle will ignore the signal.^26^ The entire system is set up as an n-body problem and integrated forward in time using the leapfrog formulas.^37,38^

All muscle attributes are initially read in from a setup file. A estimated set of muscle attributes are listed in Table 1.^39–41^ Callback functions allow the user to adjust these muscle attributes either in groups or individually at any time during the simulation. The callback functions also allow the user to ablate or excite nodes in groups or individually, change views, take snapshots, make videos, and store model settings at any time in a running simulation.

**Table 1.** Typical Parameter Run Values.

|  | Values |
| --- | --- |
| Muscle Force Per Mass Ratio, mm/ms<sup>2</sup> | 596.00 |
| Rough Estimated Mass of Left Atrium, g | 25.00 |
| Beat Period, ms | 1000.00 |
| Muscle Compression Stop Fraction | 0.70 |
| Base Muscle Contraction Duration, ms | 200.00 |
| Base Muscle Refractory Duration, ms | 200.00 |
| Base Conduction Velocity, mm/ms | 0.50 |

### Hardware and Software

The main body of the code is written in C and C++. The computationally intensive portions of the code were written in compute unified device architecture (CUDA) and offloaded to the graphics processing units (GPU) for acceleration. The nodes and vertices of the underlining atrium structure were generated in Blender using a python API to scrape all the nodes and muscle connections and store them in an input file. We then read that file into our program which builds the atrium’s internal structure. NVIDIA Nsight was used to optimize the code. The software was run on a workstation equipped with two RTX A6000 GPUs, each with over 10,000 CUDA cores and 48 GB of GDDR6 video memory. The graphical interface was written using Open Graphics Library (OpenGL).

## RESULTS

### One-Dimensional Model

Our group began work on a one-dimensional model to prove that an electrical wavefront periodically stimulated from a single node (the sinus node) would propagate down a strand of cardiac muscles and initiate a chain reaction of contractions (a beat).^33,35,42^ Turning nodes off (ablation) to see if the electrical wavefront would terminate at the off nodes was tested. Nodes were excited in front of the electrical wavefront to see if a competing wavefront would propagate from this location and interrupt the original wavefront (an ectopic event). A graphical user interface was also developed using OpenGL (Figure 1).

**Figure 1.**
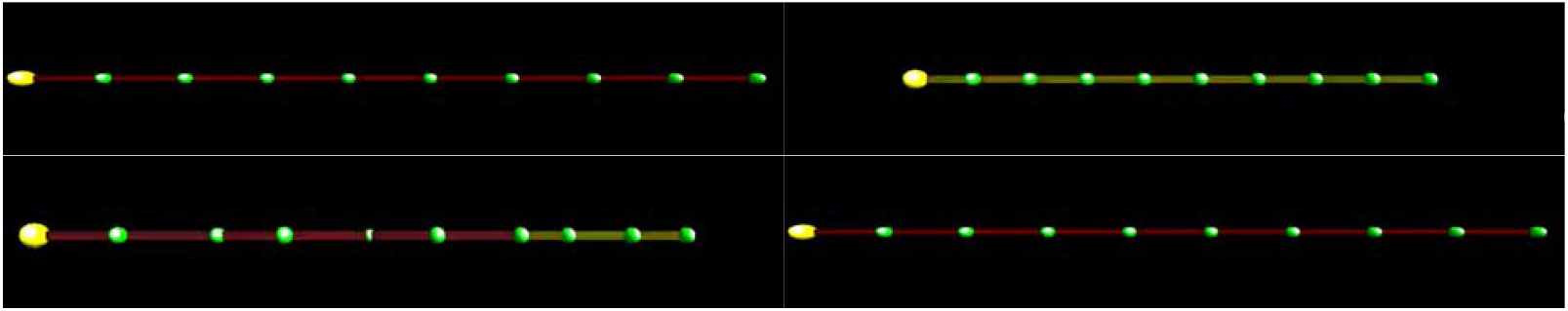
Screenshots represent the simulated cardiac muscle tissue completing a beat cycle. The large yellow sphere represents the sinus node. The smaller green spheres represent the nodes that connect muscle fibers. When the muscles are ready to contract, they are red; when contracting, they are yellow; and when recharging, they are pink.

### Two-Dimensional Model

By building on the insights obtained from the one-dimensional model, we created a more complex two-dimensional model that incorporated more sophisticated electrical pathways. To create this model, we connected the end of each muscle chain back to the stimulating node (Figure 2). This allowed us to test the propagation of electrical wavefronts in two directions and to observe how they terminated upon encountering each other. With the introduction of two dimensions, we were able to create slow and fast pathways that facilitated the production of reentry tachycardias in the model, characterized by the seizure of beat control from the stimulating node. These tachycardias were simulated by blocking certain electrical pathways, and we demonstrated that they could be eliminated through simulated ablation, further adding to our understanding of the underlying mechanisms of arrhythmias.^10,16,17^ The two-dimensional model was a critical step in advancing our understanding of arrhythmias.

**Figure 2.**
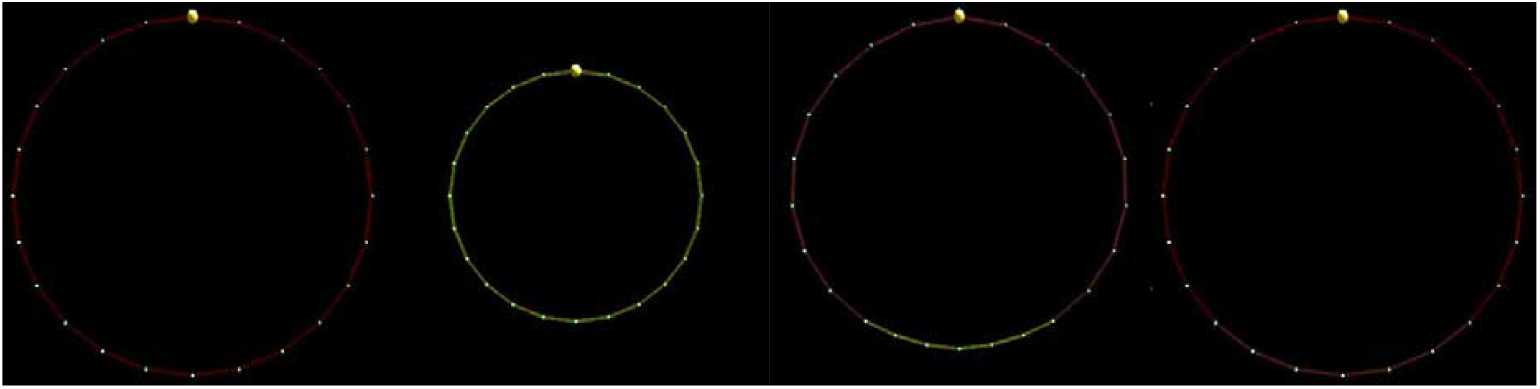
Screenshots represent a two-dimensional ring of myocardium completing a beat.

### Three-Dimensional Model

The transition to a three-dimensional model allowed us to test the propagation of electrical wavefronts from stimulated nodes in all directions, and to observe their termination on muscles that were not fully recharged or on nodes that had been ablated (Figures 3-5). The code enabled us to simulate different muscle conditions in the model, which led to the development of chaotic behavior similar to AF. This was achieved through a combination of published research and our own experimentation.^2,3,9,10,30,43^

**Figure 3.**
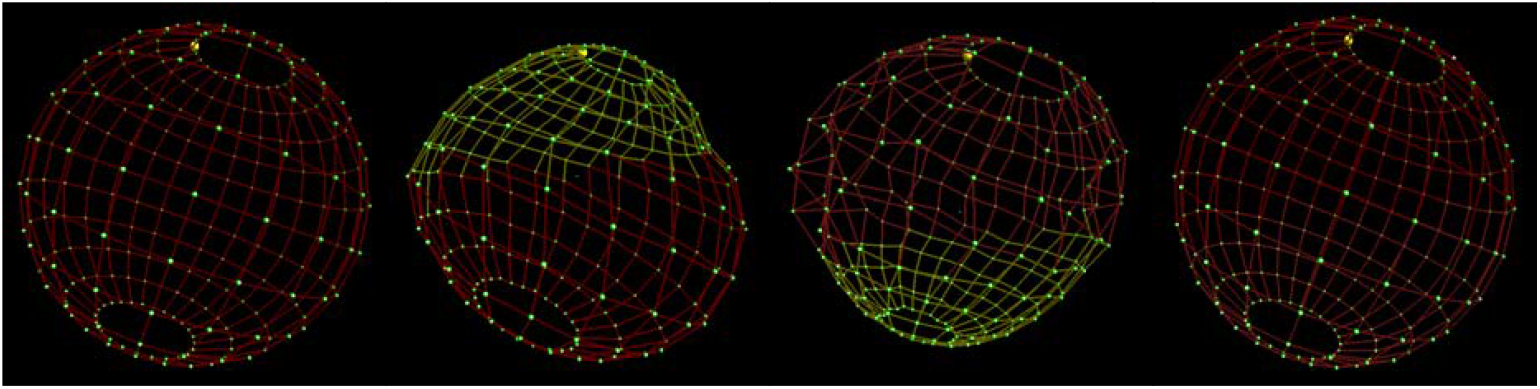
Screenshots represent the simulated atrium completing a beat.

**Figure 4.**
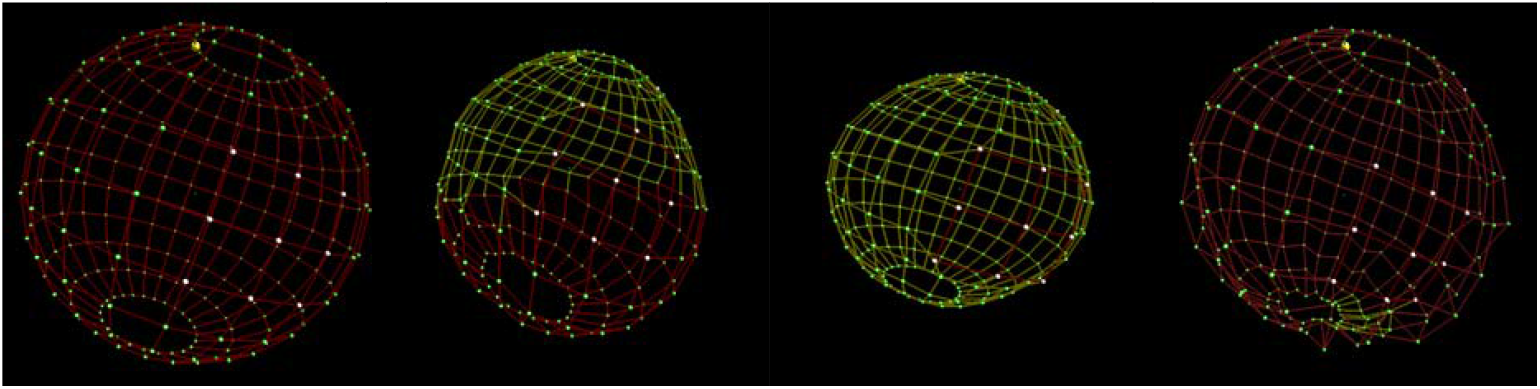
Screenshots show that the simulated ablation (white nodes) leaves this area electrically inert. The myocardium connecting ablated nodes never contracts.

**Figure 5.**
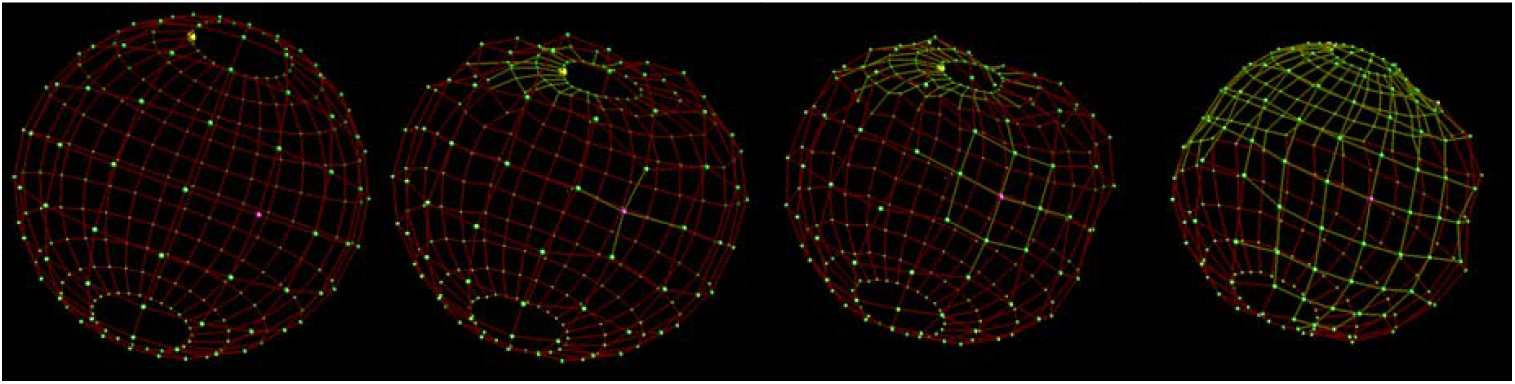
Screenshots show an ectopic signal (purple node) causing an out-of-step contraction of the myocardium, which is competing with the sinus node.

With the ability to create a functioning three-dimensional model, we were then able to perform simulations of selected node ablations in order to eliminate the chaotic behavior and restore sinus rhythm. This demonstrated the potential of the model to provide insights into the mechanisms underlying AF and make informed decisions on the development of effective therapies.^30^ The shift to a three-dimensional model also required a rewriting of our viewing and user input code to accommodate the additional dimension. This was essential for visualizing and manipulating the model in real time and allowed us to interact with the simulations in a dynamic and intuitive way.

### Parallel Processing and GPU Acceleration

The utilization of parallel processing and GPU acceleration are crucial in achieving real-time simulations in three dimensions. The increased node and muscle count made it imperative to rewrite and accelerate the code to meet computational demands (Figure 6).^44^ To achieve this, the computationally intensive portions of the code were redesigned and written in compute unified device architecture (CUDA), which enables direct access to the computing cores of modern GPUs for acceleration.^44^

**Figure 6.**
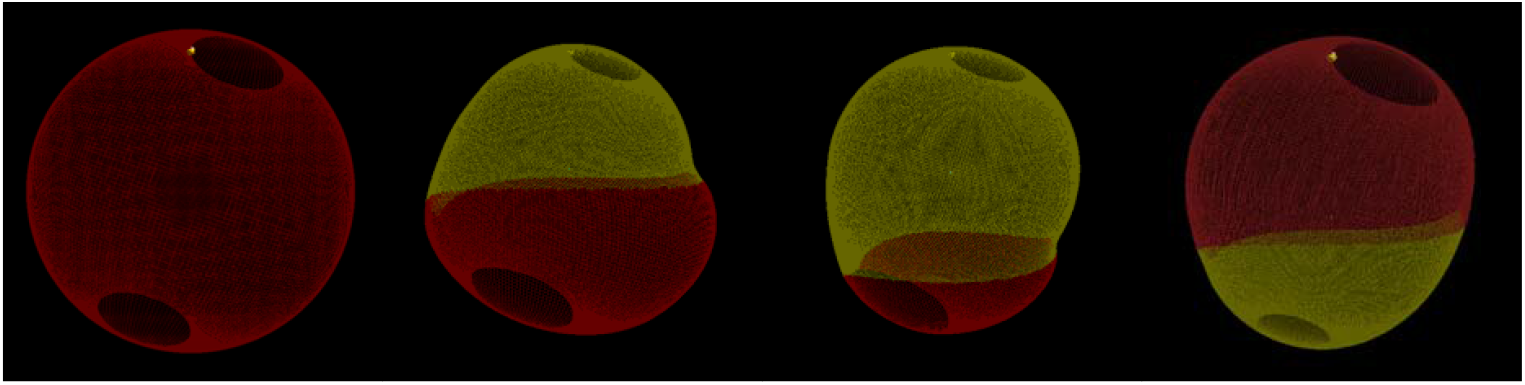
Screenshots show the more refined model with substantially more nodes and myocardium going through a contraction cycle.

The release of CUDA by NVIDIA Corporation in 2007 greatly reduced the financial barriers to available computing power for researchers. The use of supercomputers in the past was limited by their cost and size. These computers often cost tens of millions of dollars, operated in giga-flops (billion floating point operations per second), and were the size of a small room. In contrast, modern GPUs can be integrated into a laptop, cost under $1,000, and operate in tera-flops (trillion floating point operations per second). For instance, a Cray C90, which was the dominant computer in the 90s, cost around $30 million and had a performance rate of 2.3 gigaflops. Today’s NVIDIA GTX 4080, however, can be held in your hand, costs around $1,200 dollars and has a performance rate of 64 tera-flops which makes it 701 million times more cost-effective than the Cray C90. The implementation of parallel processing and GPU acceleration has greatly improved the computational capabilities and efficiency of this project, allowing for real-time simulations in three dimensions.

### Idealized Left Atrium

At this point, the model was spherical, with sections removed for the superior and inferior vena cava. This produced a crude representation of the right atrium which was sufficient for model development. The node positions and muscle connections were developed with code written entirely in-house. Most complex atrial arrhythmias spawn from the left atrium and we needed to start testing on a more anatomically accurate structure.^5,18,45^ Hence, we used Blender, an open-source 3D rendering software package, to aid in the development of the node and muscle connections of an idealized left atrium. The structure contained the four pulmonary veins in the proper locations as well as the mitral valve (see Figure 7). We removed the appendage in this model for simplification, though we knew it had to be a major component of future designs because it is where blood pools during AF causing clots that may result in strokes and heart attacks.^5,18,45,46^ Using this model we were able to take the techniques we used in the two-dimensional model and create a slowed pathway between pulmonary vein groupings that would send the model into atrial flutter.^8,11,47^ We were also able to set muscle attributes that would send the model into a chaotic web of rotational activation that closely resembled AF.^2,3,8,9^

**Figure 7.**
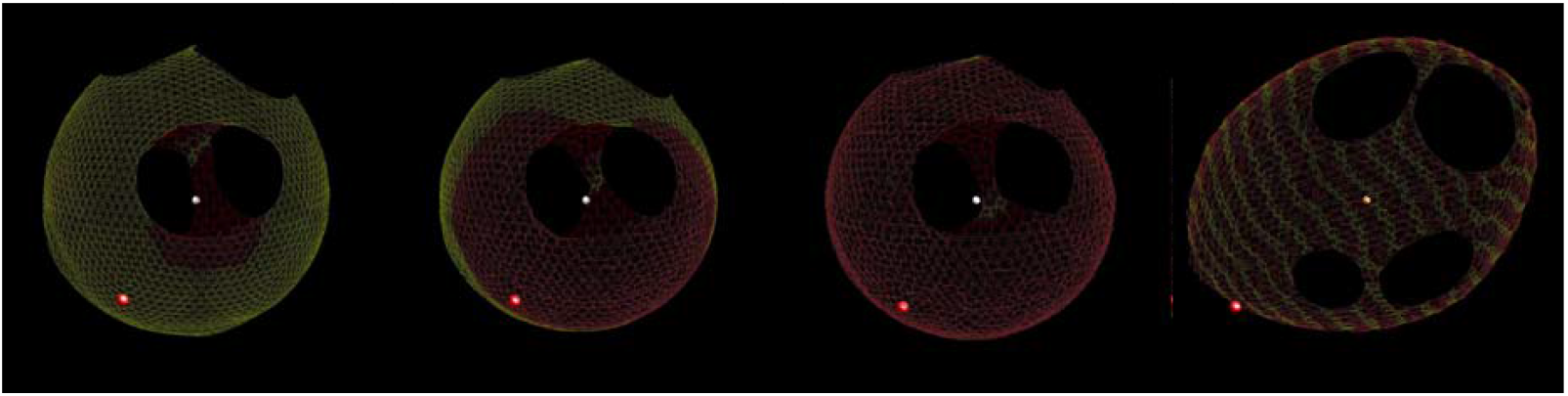
The first two screenshots are of the idealized left atrium in a normal beat. The third is a screenshot of the atrium in flutter due to the slowed pathway between the pulmonary vein cluster. The fourth screenshot is of the atrium in AF.

### Patient-Specific Models

The ultimate objective of this project is to develop patient-specific models to enable electrophysiologists (EPs) to evaluate and refine treatment strategies before performing surgery. The model can generate simulations based on any set of initial conditions (Figures 8 and 9). This capability will allow EPs and researchers to simulate known procedures and study their outcomes, as well as explore novel and untested approaches to expand their understanding and knowledge in the field.

**Figure 8.**
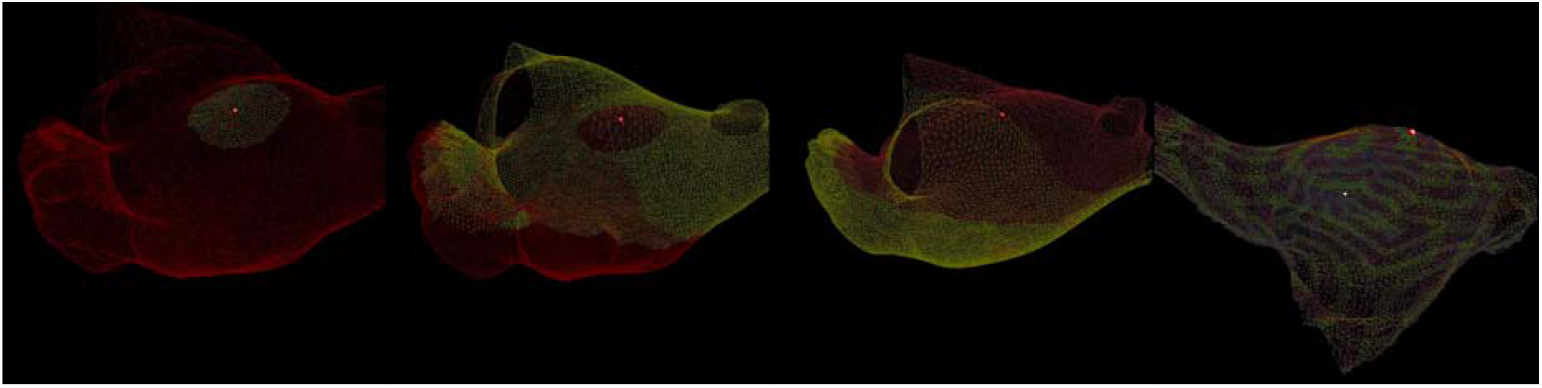
The first three screenshots of a realistic left atrium beating normally. The fourth screenshot is of the atrium in AF.

**Figure 9.**
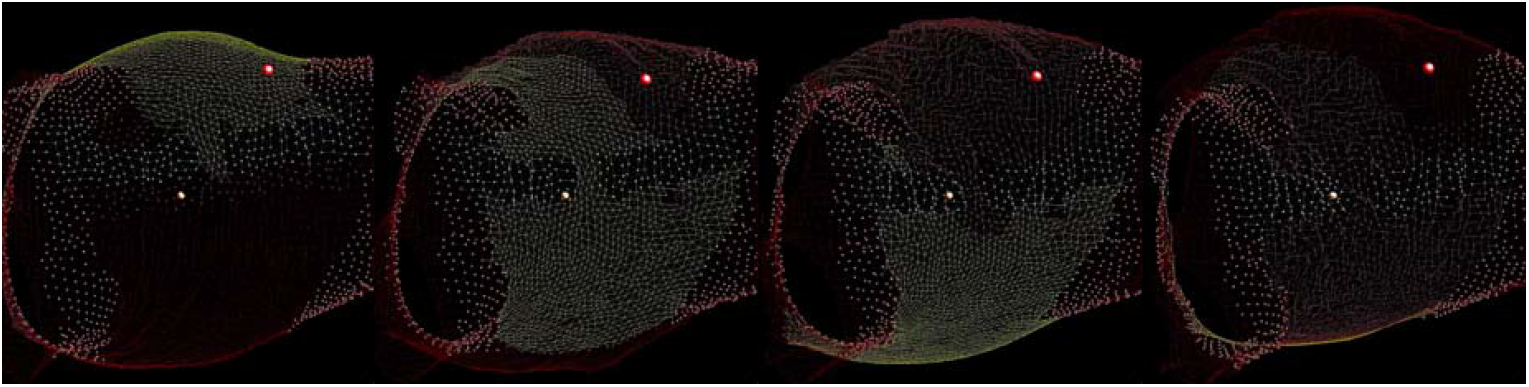
Screenshots of a realistic left atrium with appendages removed for better visualization. In this simulation, we performed pulmonary vein isolation and a faulty roof ablation. This put the model into an atrial flutter. In the running simulation, we fixed the roof ablation which put the atrium back into sinus rhythm.

## DISCUSSION

We have shown that a strand of cardiac muscles can be simulated using n-body techniques to produce a beat. This strand of simulated cardiac muscles can be used to create a semi-closed mesh of cardiac tissue in the form of the left atrium. Muscle attributes can be set in the mesh to create a substrate that will go into an arrhythmia with a properly placed ectopic signal. The model that results has demonstrated a possible mechanism for AF.^15,21^ Input data existing of shape and muscle attributes can create a model specific to that data set. We have created an interface that allows the user of the model to interact with the model in real time. The user can perform simulated ablations, introduce ectopic events, and change muscle attributes such as recharge rate and conduction velocity.

## Conclusion

The study revealed that our model has the potential to offer insights into the investigation of arrhythmias and the development of more efficient therapies. Further refinement of the model is needed; however, we are one step closer to our ultimate goal of having our model take input data from real patients and provide a model specific to that data set; providing a tool for arrhythmia specialists in their research and treatment of cardiac conditions.

## Limitations

The model’s limitations are dictated by the accuracy and availability of patient-specific data. While the computational power is sufficient to refine the model, data quality will directly impact the model’s accuracy and usefulness.

## Data Availability

No data produced

## ARTICLE INFORMATION

## Acknowledgments

We express our gratitude to Tarleton’s mathematics department for use of their high-performance computing lab.

## Media Links

https://youtube.com/shorts/pI-vvaOj4vY

https://youtu.be/WPIV6oc-JwI

https://youtube.com/shorts/oX1d83g5jVo?feature=share

https://youtube.com/shorts/YRGCbtdxYMo?feature=share

https://youtube.com/shorts/zot767ajaak?feature=share

https://youtu.be/u16OUmRAI90

https://youtu.be/T4svIVG_vb8

https://youtu.be/vXgKx36qYWY

https://youtu.be/xrgVX_XtdeE

https://youtube.com/shorts/5tqE-lyCAKg?feature=share

https://youtu.be/yQWRlOCvpe4

https://youtube.com/shorts/iEqqajYz0qM?feature=share

https://youtu.be/l4ZWjOMAp5Q

https://youtube.com/shorts/H-3Gj68CP9s?feature=share

